# Efficacy and Safety of Nicorandil for Prevention of Contrast-Induced Nephropathy in Patients Undergoing Coronary Procedures: A Systematic Review and Meta-Analysis

**DOI:** 10.1101/2024.09.15.24313706

**Authors:** Ayesha Imran Butt, Fazila Afzal, Sukaina Raza, FNU Namal, Dawood Ahmed, Hassaan Abid, Muhammad Hudaib, Zainab Safdar Ali Sarwar, Soha Bashir, Asadullah Khalid, Umer Hassan, Mohammad Ebad Ur Rehman, Huzaifa Ahmad Cheema, Ali Husnain, Usama Anwar, Muhammad Mohid Tahir, Adeel Ahmad, Wajeeh Ur Rehman, Raheel Ahmed

## Abstract

**Background:** Contrast-induced nephropathy (CIN) is a potentially serious complication of intravenous or intra-arterial contrast administration during angiographic procedures that results in renal dysfunction. This meta-analysis assesses the efficacy and safety of nicorandil for the prevention of CIN in patients undergoing percutaneous coronary intervention (PCI) or coronary angiography (CAG).

**Methods:** Cochrane Central Register of Controlled Trials, MEDLINE, Embase, and ClinicalTrials.gov were used to perform a thorough literature search from their inception to July 2024. A random-effects meta-analysis was performed on RevMan and pooled estimates were presented as forest plots. The Mantel-Haenszel method was used for dichotomous outcomes and risk ratios (RRs) were calculated along with 95% confidence intervals (95% CI).

**Results:** This meta-analysis included 12 RCTs consisting of 2787 participants (nicorandil: 1418, control: 1394). The use of nicorandil was protective against CIN (RR 0.38, 95% CI 0.29-0.50). The incidence of major adverse events was comparable in both groups (RR 0.77, 95% CI 0.52-1.13, p=0.18). Similarly, the use of nicorandil did not affect the risk of developing stroke (RR 1.05), myocardial infarction (RR 0.90), heart failure (RR 0.81), cardiac death (RR 0.90), and dialysis (RR 0.70).

**Conclusion:** This study revealed that nicorandil effectively reduced the risk of developing CIN in patients undergoing angiographic procedures like PCI or coronary angiography. However, more RCTs should be conducted for a more definitive conclusion.

## Introduction

Percutaneous coronary intervention (PCI) is defined as any procedure that is performed to widen the lumen of an obstructed coronary artery and involves passing a catheter through the skin and into a blood vessel to the site of obstruction so the blockage can be compressed or removed. PCI is the first-line treatment for coronary artery disease, one of the leading causes of death globally. Coronary angiography (CAG) is a diagnostic procedure involving the injection of contrast dye to visualize blood flow through the heart. Contrast-induced nephropathy (CIN) is a well-known complication of PCI and CAG. CIN is defined as a rise in serum creatinine of at least 0.5 mg/dL or a 25% increase from baseline within 48 to 72 hours after contrast exposure.^1^ The incidence of CIN is increasing due to the introduction of new procedures requiring contrast injection. The incidence of CIN following PCI ranges from 2-20% depending upon the baseline kidney function of the patient. It can be even higher in an emergency clinical setting.^2^ CIN after PCI is significantly associated with a higher likelihood of extended hospital stays as well as an increased risk of complications like death, myocardial infarction (MI), bleeding, and recurrent renal injury after discharge.^3^ Patients who develop post-PCI CIN are at more than threefold greater risk of all-cause mortality compared to the patients who do not experience CIN after PCI.^4^

Investigations have shown that the pathogenesis of CIN following PCI and CAG is quite complex and multifactorial, but ischemia and hypoxia of renal cells due to low blood flow to the kidney may play an important role in the pathogenesis.^5^ Nicorandil works by nitric oxide (NO) donation and K-ATP channel opening, which leads to the relaxation of smooth muscle cells of the vessels causing vasodilation. Nicorandil may help prevent hypoxia and ischemia of the renal cells by maintaining blood supply to the renal cells via its vasodilatory effects.^6^ Some animal studies confirm that K-ATP channel openers, including nicorandil, prevent nephropathy related to ischemia or hypoxic injury.^7,8^ Randomized controlled trials (RCTs) conducted over time have shown conflicting results, but a few have shown a trend of improved outcomes related to CIN with nicorandil following PCI and CAG. Although there is some promising evidence supporting the use of nicorandil, its inclusion in the standard PCI/CAG protocols remains debated. Nicorandil has been approved as a long-term therapy for stable angina in Europe and Japan,^9^ but according to the 2021 ACC/AHA/SCAI Guideline for Coronary Artery Revascularization, nicorandil is not currently recommended for use during PCI or CAG.^10^

Several meta-analyses have assessed the use of nicorandil for the prevention of CIN due to PCI/ CAG.^11–13^ The results were promising as they showed a statistically significant reduction in the cases of CIN for patients given nicorandil as compared to the patients who did not receive any preventive medicine for CIN during the procedure. However, several new RCTs have been published in recent years. A pooled analysis may help establish guidelines for CIN prevention following PCI/CAG. In addition, the safety profile of nicorandil can be clarified. Therefore, we have undertaken a systematic review and meta-analysis to assess the efficacy and safety of nicorandil for the prevention of post-PCI/CAG CIN.

## Materials and Methods

Our meta-analysis was conducted according to the guidelines provided in the Cochrane Handbook for Systematic Reviews of Interventions and reported according to the Preferred Reporting Items for Systematic Reviews and Meta-Analysis (PRISMA) statement^14,15^. Our protocol is registered with PROSPERO (CRD42024572031).

### Data Sources and Searches

Cochrane Central Register of Controlled Trials, MEDLINE, Embase, and ClinicalTrials.gov were used to perform a thorough electronic search from their inception to July 2024. Reference lists of included studies and similar systematic reviews were also screened to identify further pertinent studies.

The following terms (“Nicorandil”) AND (“Coronary Angiography” OR “Percutaneous Coronary Intervention”) were used as either Medical Subject Heading (MeSH) terms or keywords.

### Eligibility Criteria

All RCTs that compared the use of nicorandil in the prevention of CIN among patients undergoing PCI/CAG to a placebo or standard treatment were included in this meta-analysis.

Parameters for exclusion included any study design other than RCTs such as quasi-randomized trials observational studies, and studies conducted on animals. No language or date restrictions were applied.

### Study Selection and Data Extraction

We used Rayyan to narrow down and remove duplicates of all the articles yielded by our online search. Two authors (A.I. and S.R.) independently performed a screening of titles and abstracts to exclude all irrelevant articles. Full-text screening in accordance with our eligibility criteria was then performed on the remaining studies. Any discrepancies over the selection of studies were settled by a third author (M.E.U.R).

Relevant data was extracted into a pre-piloted Excel spreadsheet which included author name, publication year, country name, sample size, study design, mean age, gender, hypertension, diabetes, smoking, cardiac procedure, nicorandil regimen (dose, duration, frequency, and route) and outcomes.

### Outcomes

The primary outcome of our study was CIN while the secondary outcomes were major adverse effects, MI, cardiac death, heart failure, stroke, and dialysis.

### Risk of Bias Assessment

The risk of bias in the included studies was evaluated using the revised Cochrane Risk of Bias tool for randomized trials (RoB 2.0)^16^. This tool assesses bias in five domains which comprises of: (1) bias caused by the randomization process; (2) bias due to deviations from intended interventions; (3) bias arising from missing outcome data; (4) bias in the measurement of the outcome, and (5) bias in the selection of the reported result. The risk of bias for each included study was assessed by two investigators (D.A. and F.A.), with high, low, or some concerns of bias being reported. Any disagreement was settled by a senior investigator (M.E.U.R).

### Data Synthesis

To conduct the meta-analyses, we used Review Manager (RevMan, Version 5.4.1) software. The Mantel-Haenszel method was used for dichotomous outcomes. Risk ratios (RRs) and corresponding 95% confidence interval (CI) were extracted. A random-effects model was used to carry out the meta-analyses. Pooled estimates were presented as a forest plot and to assess statistical heterogeneity, the Higgins I-square statistic was calculated.

## Results

### Search Results

The initial search garnered 798 articles. Title and abstract screening resulted in the exclusion of 300 articles, and 236 out of the remaining 248 studies were excluded after full-text screening. Finally, 12 articles were included in this meta-analysis. The detailed screening process is depicted in Figure 1.

**Figure 1:**
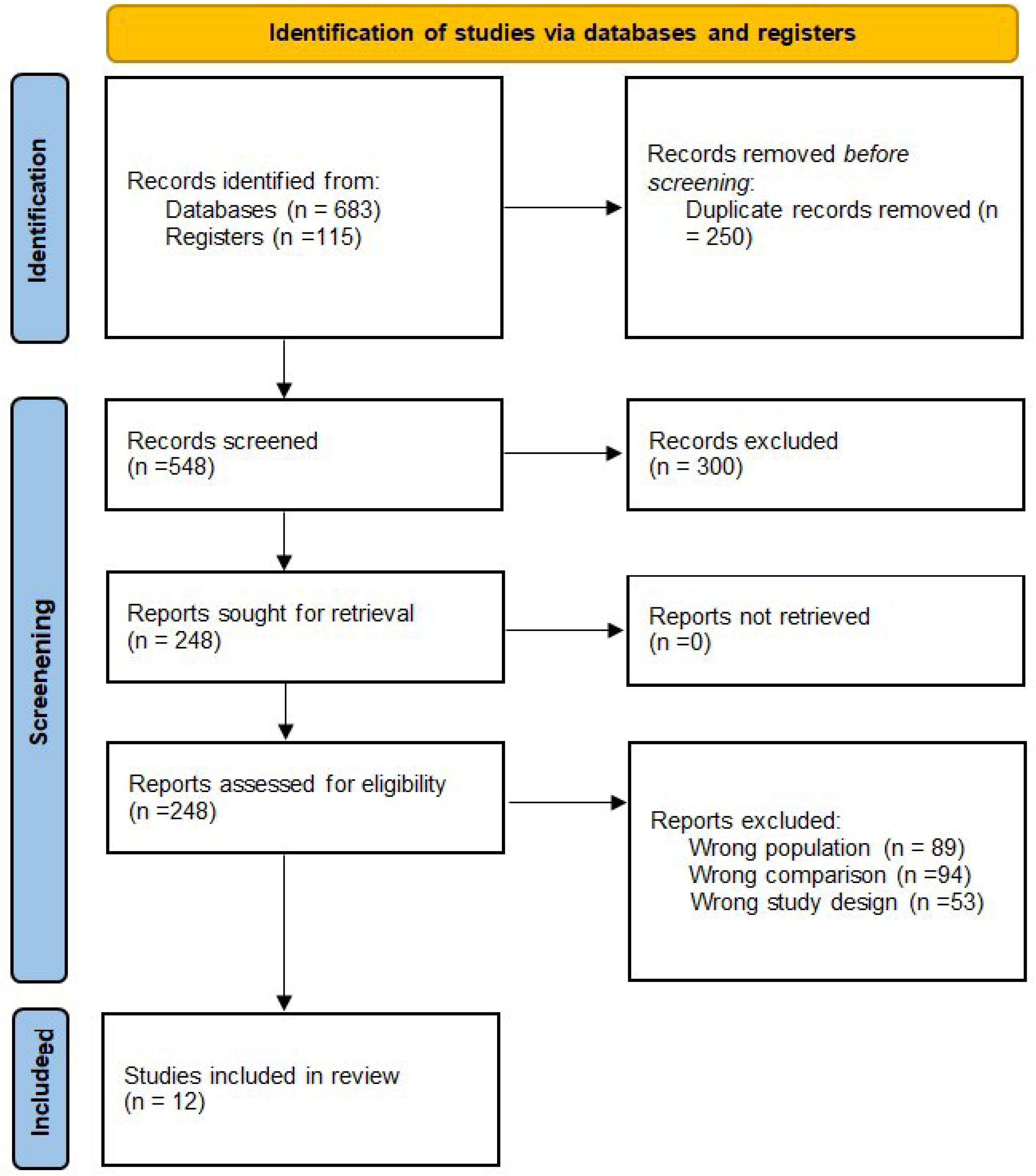
PRISMA Flowchart

### Study Characteristics

This meta-analysis included 12 RCTs that met the eligibility criteria.^17–28^ A total of 2787 patients undergoing any cardiac procedure involving contrast were included. In these 12 RCTs conducted across 6 countries, 1418 patients received nicorandil (IV nicorandil =256, oral nicorandil =1162) and 1369 patients were assigned to the control group. Oral nicorandil was used in 9 studies whereas IV nicorandil was used in 3 studies. The publication years ranged from 2013 to 2024. Study characteristics are reported in Table 1.

**Table 1:**
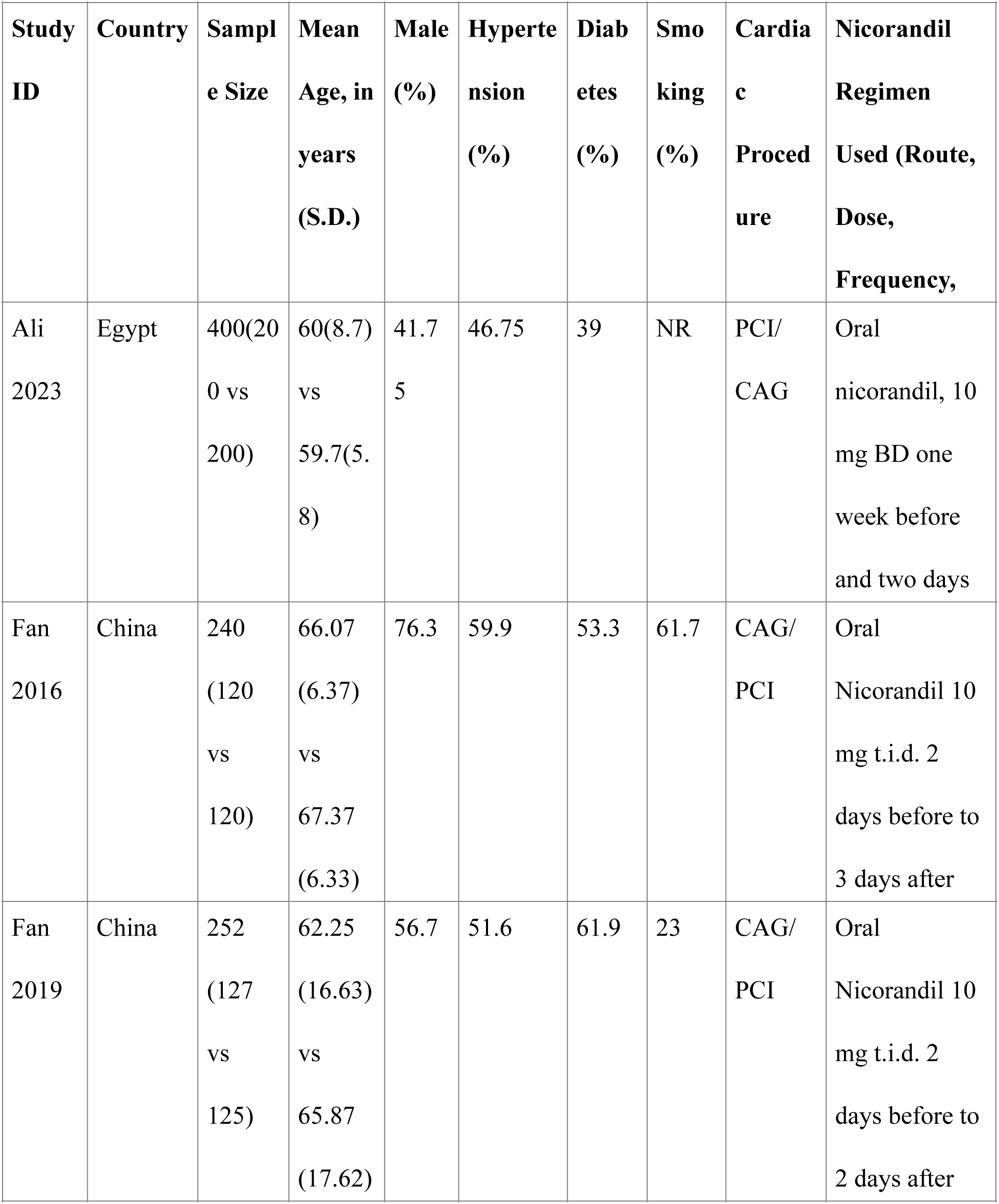
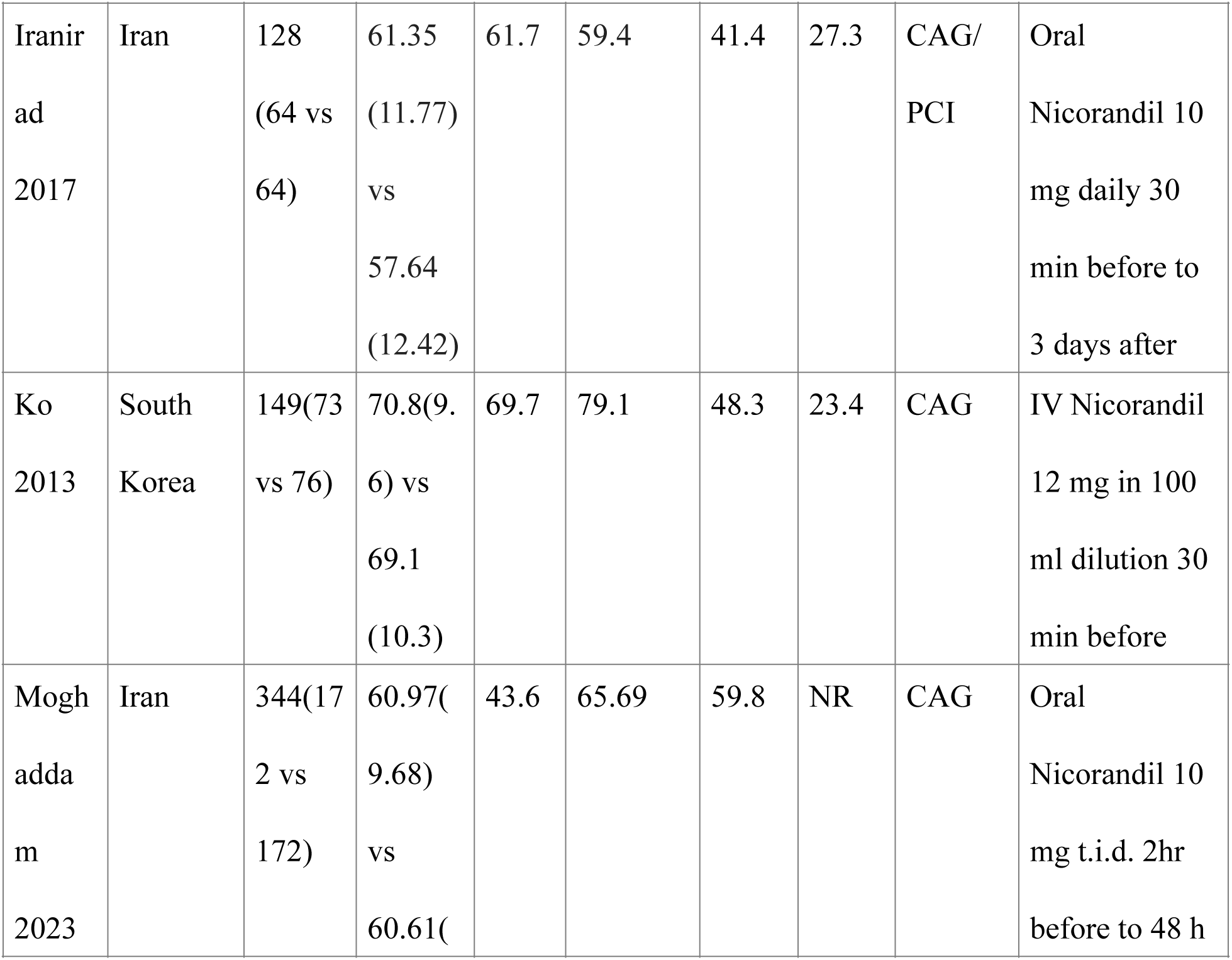
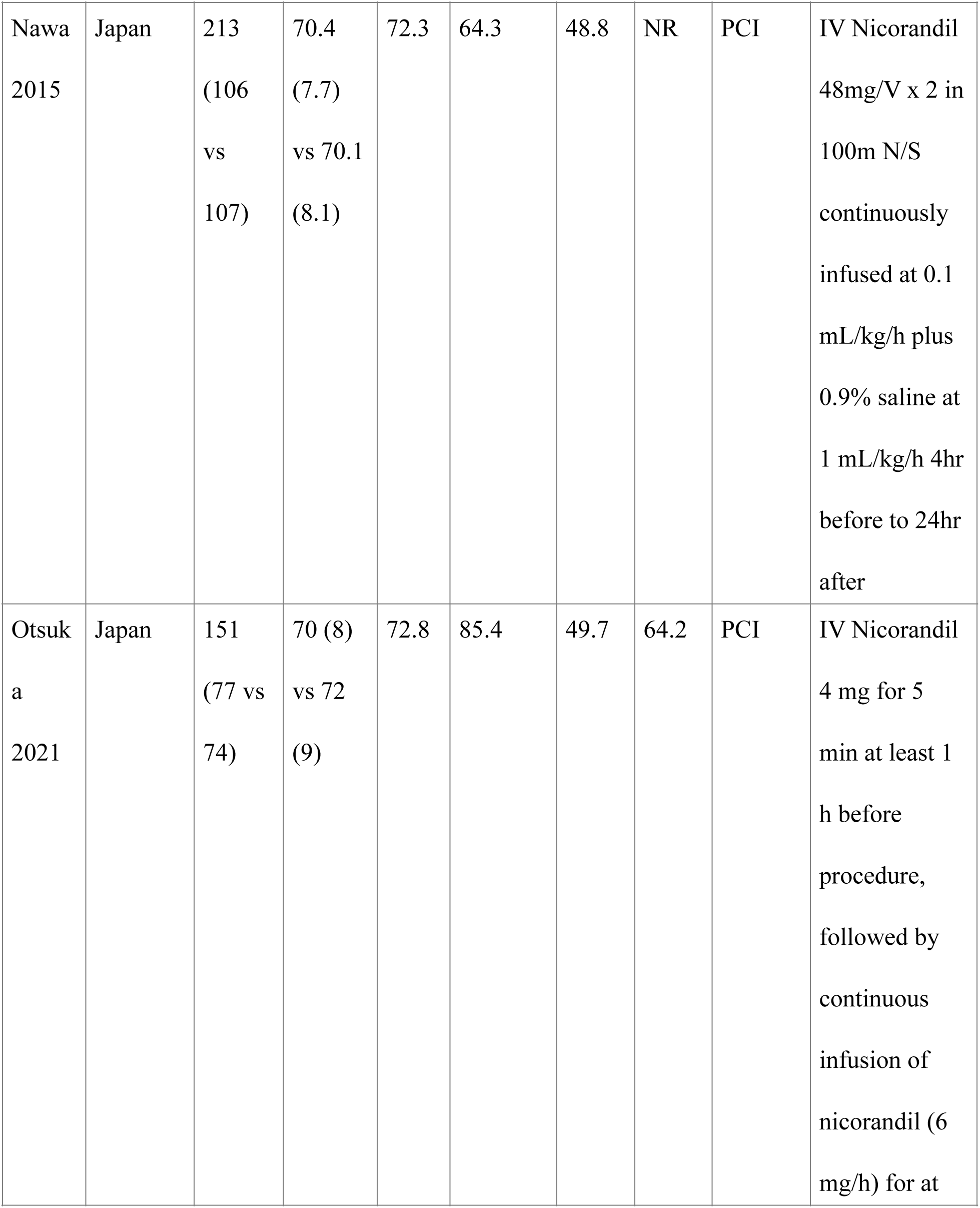
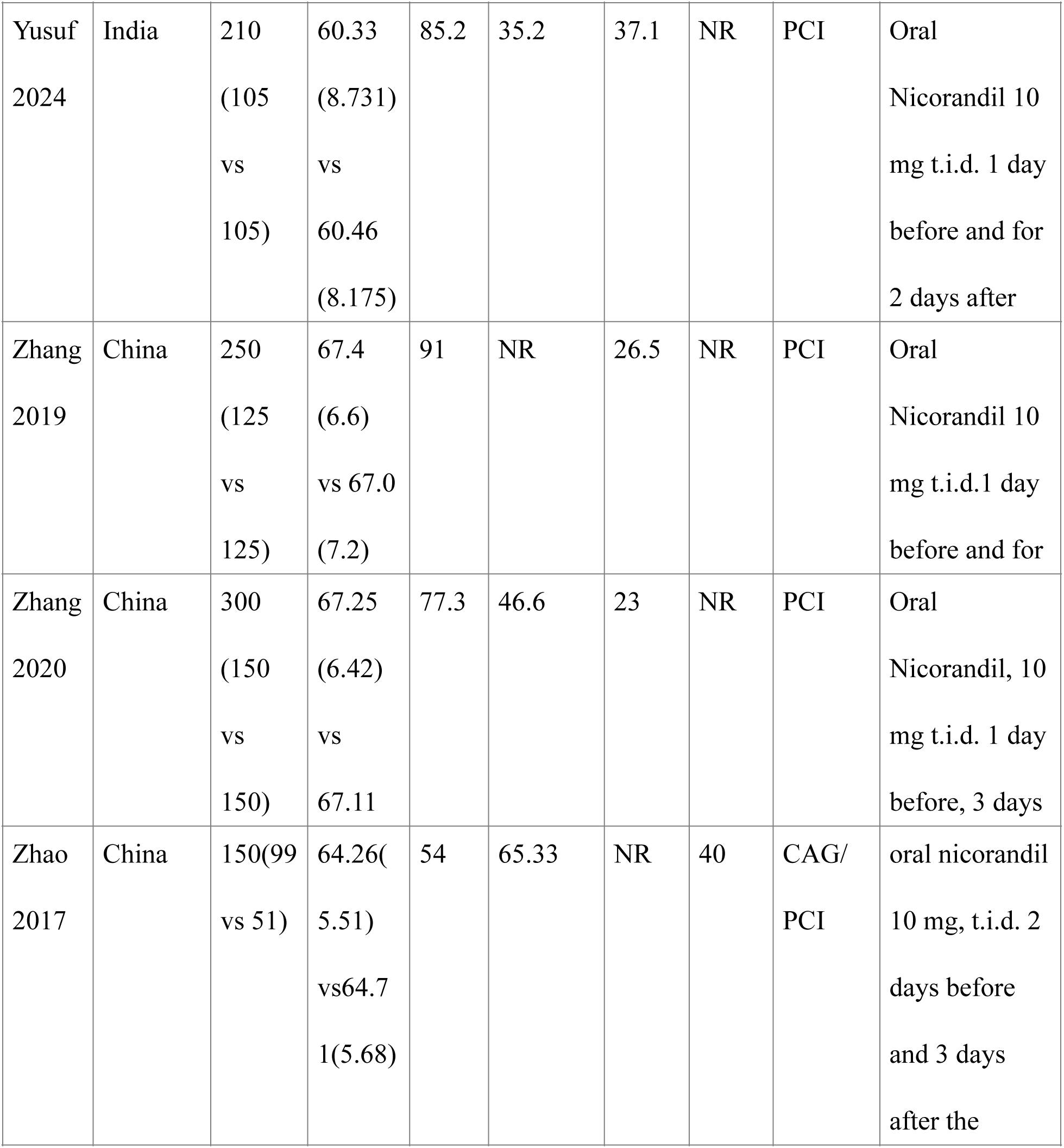
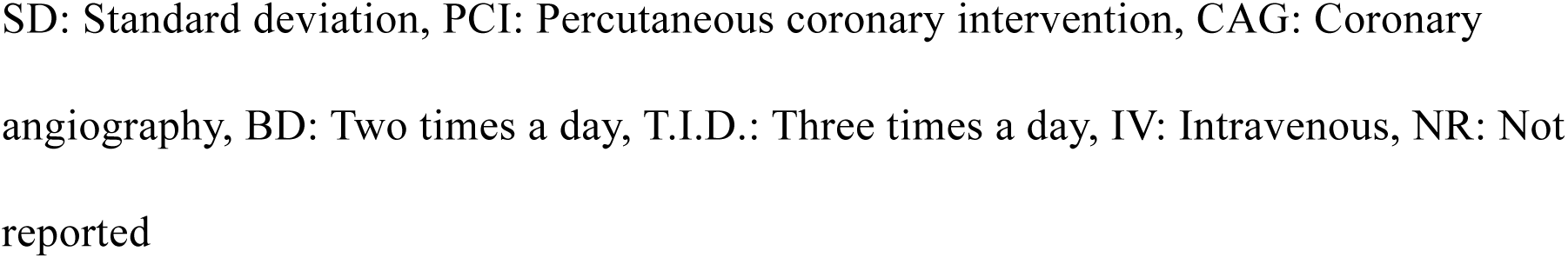
Characteristics of Included Studies.

| **Study ID** | **Country** | **Sample Size** | **Mean Age, in years (S.D.)** | **Male (%)** | **Hypertension (%)** | **Diabetes (%)** | **Smoking (%)** | **Cardiac Procedure** | **Nicorandil Regimen Used (Route, Dose, Frequency,** |
| --- | --- | --- | --- | --- | --- | --- | --- | --- | --- |
| Ali 2023 | Egypt | 400(200 vs 200) | 60(8.7) vs 59.7(5.8) | 41.75 | 46.75 | 39 | NR | PCI/CAG | Oral nicorandil, 10 mg BD one week before and two days |
| Fan 2016 | China | 240 (120 vs 120) | 66.07 (6.37) vs 67.37 (6.33) | 76.3 | 59.9 | 53.3 | 61.7 | CAG/PCI | Oral Nicorandil 10 mg t.i.d. 2 days before to 3 days after |
| Fan 2019 | China | 252 (127 vs 125) | 62.25 (16.63) vs 65.87 (17.62) | 56.7 | 51.6 | 61.9 | 23 | CAG/PCI | Oral Nicorandil 10 mg t.i.d. 2 days before to 2 days after |
| Iranir<br>ad<br>2017 | Iran | 128<br>(64 vs<br>64) | 61.35<br>(11.77)<br>vs<br>57.64<br>(12.42) | 61.7 | 59.4 | 41.4 | 27.3 | CAG/<br>PCI | Oral<br>Nicorandil 10<br>mg daily 30<br>min before to<br>3 days after |
| Ko<br>2013 | South<br>Korea | 149(73<br>vs 76) | 70.8(9.<br>6) vs<br>69.1<br>(10.3) | 69.7 | 79.1 | 48.3 | 23.4 | CAG | IV Nicorandil<br>12 mg in 100<br>ml dilution 30<br>min before |
| Mogh<br>adda<br>m<br>2023 | Iran | 344(17<br>2 vs<br>172) | 60.97(<br>9.68)<br>vs<br>60.61( | 43.6 | 65.69 | 59.8 | NR | CAG | Oral<br>Nicorandil 10<br>mg t.i.d. 2hr<br>before to 48 h |
| Nawa<br>2015 | Japan | 213<br>(106<br>vs<br>107) | 70.4<br>(7.7)<br>vs 70.1<br>(8.1) | 72.3 | 64.3 | 48.8 | NR | PCI | IV Nicorandil<br>48mg/V x 2 in<br>100m N/S<br>continuously<br>infused at 0.1<br>mL/kg/h plus<br>0.9% saline at<br>1 mL/kg/h 4hr<br>before to 24hr<br>after |
| Otsuk<br>a<br>2021 | Japan | 151<br>(77 vs<br>74) | 70 (8)<br>vs 72<br>(9) | 72.8 | 85.4 | 49.7 | 64.2 | PCI | IV Nicorandil<br>4 mg for 5<br>min at least 1<br>h before<br>procedure,<br>followed by<br>continuous<br>infusion of<br>nicorandil (6<br>mg/h) for at |
| Yusuf<br>2024 | India | 210<br>(105<br>vs<br>105) | 60.33<br>(8.731)<br>vs<br>60.46<br>(8.175) | 85.2 | 35.2 | 37.1 | NR | PCI | Oral<br>Nicorandil 10<br>mg t.i.d. 1 day<br>before and for<br>2 days after |
| Zhang<br>2019 | China | 250<br>(125<br>vs<br>125) | 67.4<br>(6.6)<br>vs 67.0<br>(7.2) | 91 | NR | 26.5 | NR | PCI | Oral<br>Nicorandil 10<br>mg t.i.d.1 day<br>before and for |
| Zhang<br>2020 | China | 300<br>(150<br>vs<br>150) | 67.25<br>(6.42)<br>vs<br>67.11 | 77.3 | 46.6 | 23 | NR | PCI | Oral<br>Nicorandil, 10<br>mg t.i.d. 1 day<br>before, 3 days |
| Zhao<br>2017 | China | 150(99<br>vs 51) | 64.26(<br>5.51)<br>vs64.7<br>1(5.68) | 54 | 65.33 | NR | 40 | CAG/<br>PCI | oral nicorandil<br>10 mg, t.i.d. 2<br>days before<br>and 3 days<br>after the |

SD: Standard deviation, PCI: Percutaneous coronary intervention, CAG: Coronary angiography, BD: Two times a day, T.I.D.: Three times a day, IV: Intravenous, NR: Not reported

### Risk of Bias in Included Studies

The quality assessment of the included studies is presented in Supplementary Figure 1. Overall, four studies were judged to be at low risk of bias while two studies were at high risk of bias due to issues in the randomization process, measurement of the outcome, and reporting of the results. An inadequate randomization process, deviations from the intended interventions, and flawed measurement of the outcome led to some concerns of bias in six studies.

### Meta-Analysis of Primary Outcome: Contrast-Induced Nephropathy

Twelve studies were included in the analysis of CIN, involving a total of 2787 patients (1418 Nicorandil vs 1369 Control). The analysis revealed that nicorandil significantly reduced the incidence of CIN as compared to the control group (RR: 0.38, 95% CI: 0.29-0.50, p < 0.00001, I^2^ = 0%) (Figure 2). In the subgroup analysis for the route of administration, the use of nicorandil through oral route revealed statistically significant reduction in incidence of CIN (RR: 0.36, 95% CL: 0.27-0.49, P < 0.00001, I^2^ = 0%) (Figure 3), whereas use of nicorandil through IV route was not significantly associated with reduction in incidence of CIN (RR: 0.46, 95% CI: 0.20-1.09, p < 0.08, I^2^ = 29%) (Figure 3).

**Figure 2:**
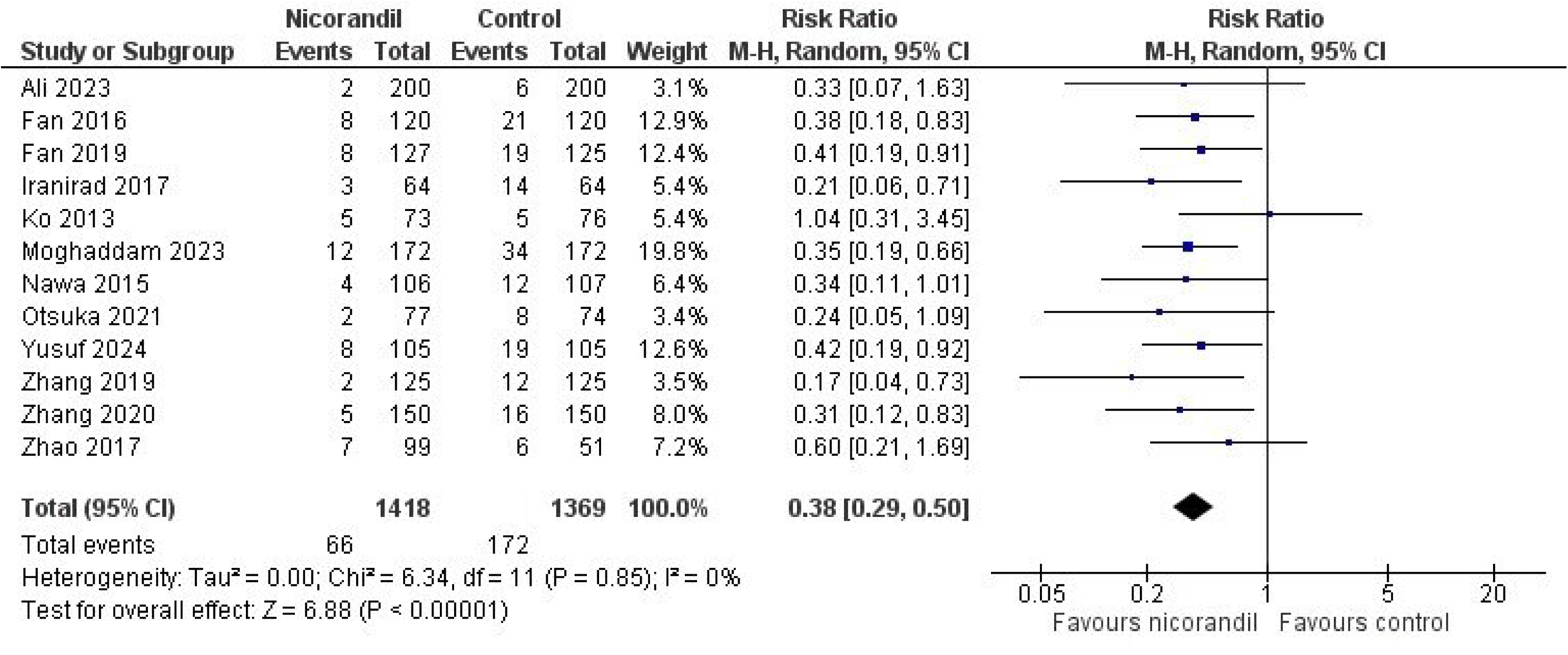
Forest Plot of Contrast Induced Nephropathy

**Figure 3:**
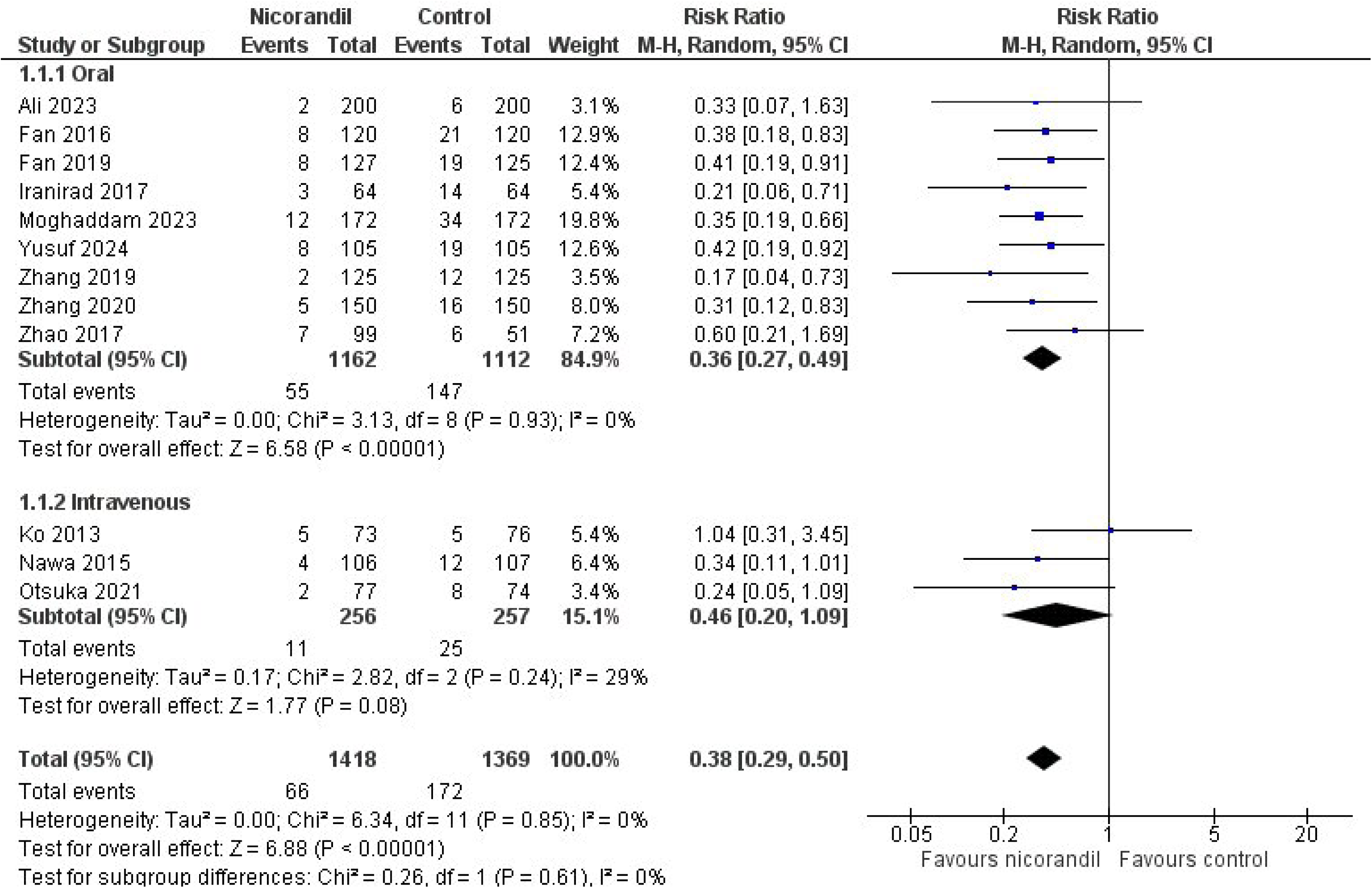
Subgroup Analysis of Contrast Induced Nephropathy for Route of Administration

### Meta-Analysis of Secondary Outcome

#### Major Adverse Events

This outcome was reported by 6 studies. The total number of patients was 1535 (767 Nicorandil vs 768 Control). According to the analysis, no statistically significant difference in major adverse events was observed (RR: 0.77, 95% CI: 0.52-1.13, p < 0.18, I^2^ = 4%) (Supplementary Figure 2).

#### Myocardial Infarction

Five studies reported this outcome. The total number of patients was 1101 (550 Nicorandil vs 551 Control). The analysis showed comparable results (RR: 0.90, 95% CI: 0.56-1.43, p < 0.65, I^2^ = 0%) between the two groups (Supplementary Figure 3).

#### Cardiac Death

Five studies reported cardiac death. The total number of patients was 1101 (550 Nicorandil vs 551 Control). The results were not statistically significant for this outcome (RR: 0.90, 95% CI: 0.28-2.88, p < 0.86, I^2^ = 0%) (Supplementary Figure 4).

#### Heart Failure

Heart failure was reported in five studies. The total number of patients was 1252 (627 Nicorandil vs 625 Control). According to the analysis, there is no statistically significant association between increased risk of heart failure and the use of nicorandil (RR: 0.81, 95% CI: 0.39-1.68, p < 0.58, I^2^ = 0%) (Supplementary Figure 5).

#### Stroke

This outcome was reported by four studies. The total number of patients was 1042 (522 Nicorandil vs 520 Control). The analysis showed comparable results (RR: 1.05, 95% CI: 0.38-2.95 p < 0.92, I^2^ = 0%) between the two groups (Supplementary Figure 6).

#### Dialysis

The analysis of three studies that reported this outcome revealed no statistically significant association between the use of nicorandil and dialysis (RR: 0.70, 95% CI: 0.11-4.44 p < 0.71, I^2^ = 0%) (Supplementary Figure 7).

## Discussion

Our meta-analysis of 12 RCTs with 2787 participants showed that nicorandil significantly reduces the risk of CIN, with no significant increase in major adverse events, MI, cardiac death, heart failure, stroke, and dialysis between the two groups.

Our meta-analysis demonstrates that nicorandil reduces the risk of CIN by approximately one-third. Previous studies support this finding. For instance, Pranata et al. analyzed seven RCTs with 1,532 subjects who underwent CAG and/or PCI. They found that nicorandil significantly decreased CIN risk (OR 0.31, p < 0.001).^29^ Another analysis by Zhan et al., which reviewed five studies with 805 patients undergoing CAG or PCI, also reported a significant reduction in CIN with nicorandil (RR 0.37, p = 0.0001).^30^ Yiliang Li’s meta-analysis of 14 RCTs involving 1,947 patients with stable angina undergoing elective PCI showed a comparable reduction (RR 0.36, p < 0.00001).^31^ In addition, Shuang Li’s meta-analysis of four RCTs with 730 patients demonstrated a 64% reduction in CIN risk with nicorandil (RR 0.36, 95% CI 0.22-0.61).^32^ Xiaofen Ma’s meta-analysis, which included 709 patients from four RCTs, supported these results by showing a reduction in CIN risk (RR 0.38, p = 0.005)^33^. Lastly, Bin Yi et al.’s pooled analysis of six RCTs with 1,229 patients undergoing elective PCI found that nicorandil lowered CIN risk (OR 0.26, p < 0.00001). Collectively, these studies reinforce our findings, highlighting the effectiveness of nicorandil in reducing CIN risk^34^.

CIN is the third major complication post-PCI, after stent restenosis and thrombosis.^35^ Its incidence varies from 0% to 24% and specifically from 3% to 14% in PCI patients, with higher rates in those with impaired renal function.^29,36^ Over 50% of high-risk patients develop CIN after PCI.^37^ While less than 5% of the general population experiences CI-AKI after coronary angiography, the risk is much higher for patients with kidney issues, and acute myocardial infarction (AMI), especially STEMI, shock, or diabetes. CI-AKI can lead to increased need for dialysis, higher rates of major cardiovascular events, longer hospital stays, and greater costs, underscoring the importance of effective prevention and treatment.^38^ The exact mechanism is unclear but may involve renal hemodynamic changes, medullary ischemia, tubular cell toxicity, reduced nitric oxide, calcium overload, apoptosis, increased blood viscosity, immune/ inflammatory response, and oxidative stress.^39–41^

Nicorandil promotes vasodilation by activating K-ATP channels, which leads to membrane hyperpolarization and reduced Ca^2+^ influx. Additionally, its nitrate-like effect generates nitric oxide, increasing cyclic GMP and activating protein kinase G, further reducing vascular resistance and enhancing vasodilation.^42–44^ Nicorandil may protect the kidneys by improving blood flow in both coronary and peripheral arteries, reducing heart strain, and improving kidney blood flow.^43,44^ Studies indicate that nicorandil may offer extra kidney protection and decrease the risk of CIN during coronary procedures likely via mechanisms such as reducing blood vessel and kidney dysfunction or by counteracting adverse effects by normalizing the TLR4/MAPK P38/NFκ-B inflammatory pathway.^45^ Research has shown that activation of K-ATP channels reduces reactive oxygen species (ROS) accumulation in renal tissue following ischemia-reperfusion injury, which helps mitigate kidney damage.^46^ Nicorandil also facilitates ischemic preconditioning, enhancing tissue resistance to ischemia. Remote ischemic preconditioning (RIPC) has been demonstrated to lower CIN risk in patients undergoing percutaneous coronary intervention, especially those at higher risk for CIN.^47^ These mechanisms may explain how nicorandil reduces CIN in patients undergoing cardiac catheterization.

Our study indicated that the oral route seemed to be more effective than intravenous (IV) nicorandil. The study by Bin Yi et al. demonstrated a consistent effect of nicorandil on CIN after elective PCI, regardless of whether it was administered orally or intravenously.^34^ Conversely, the study by Pranata et al. showed that oral nicorandil was more effective in preventing CIN than the IV form.^29^ A possible reason behind this IV inefficacy could be explained by the fact that contrast media are not metabolized in the human body but are rapidly eliminated through glomerular filtration by the kidneys. The time required to clear half of the contrast medium from the blood, known as the elimination half-life, is approximately 1–2 hours. In individuals with normal renal function, nearly 100% of the contrast medium is excreted within the first 24 hours after administration. However, in patients with impaired renal function, the elimination half-life can extend to 40 hours or more.^39^ Only Ko, Nawa, and Otsuka administered IV nicorandil.^48–50^ Ko administered it just 30 minutes prior to the procedure.^48^ Nawa provided IV nicorandil for four hours before to 24 hours after the procedure^49^. Otsuka gave nicorandil for 5 minutes at least 1 hour before the procedure, followed by continuous infusion for at least 8 hours, which was stopped at the discretion of the attending doctor^50^. In contrast, most of the other eligible RCTs administered oral nicorandil at least 2 to 7 days before the procedure and continued for 2 to 3 days afterward. Only one RCT administered oral nicorandil 30 minutes before the procedure and continued for 3 days afterward.^51^ Nicorandil has a short half-life of around 1 hour, which might result in its removal from the blood earlier than the contrast media.^52^ This could potentially explain the superior efficacy of the oral route and highlights the need for further research.

Our meta-analysis found no significant difference in major adverse events between the nicorandil and non-nicorandil groups. This is consistent with prior studies, such as Xiaofen Ma et al., which reported no significant effect of nicorandil on major adverse events (RR: 0.90, 95% CI: 0.36– 2.28, P = 0.82).^33^ Similarly, Pranata et al. found no significant association between nicorandil and major adverse events (OR: 0.67, 95% CI: 0.42–1.07, P = 0.09).^29^ Li et al. also showed no significant reduction in major adverse events with nicorandil (OR: 0.75, 95% CI: 0.49–1.15, P = 0.19), though they noted a significant reduction in postoperative MI in the nicorandil group (RR: 0.58, P = 0.001).^31^ The meta-analysis by Bin Yi found that data revealed no significant differences between the groups for mortality (OR: 0.82, P = 0.79), heart failure (OR: 0.79, P = 0.60), recurrent MI (OR: 0.35, P = 0.35), stroke (OR: 3.02, P = 0.34), or renal replacement therapy (OR: 0.47, P = 0.51) following elective PCI.^34^ In Zuo-Zhong Yu’s retrospective control study, there was no significant difference in major adverse events between the two groups within six months after PCI.^38^ On the contrary, Ishii et al. found that oral nicorandil might decrease cardiac events following PCI in dialysis patients.^53^ Nicorandil’s arterial and venous dilation effects reduce afterload and preload, respectively, thereby reducing myocardial ischemia.^44^ Remote ischemic preconditioning (RIPC) is also effective in decreasing CIN and major adverse events in patients undergoing PCI, particularly those at high risk for CIN.^35^

Despite our study showing no effect of nicorandil on heart failure, it is known that nicorandil can improve heart function in heart failure by widening coronary arteries and increasing blood flow to the heart. It also protects heart cell mitochondria and prevents heart cell death.^45^ Nicorandil’s pharmacological actions, such as ischemic preconditioning, stabilization of coronary plaques, and enhancement of impaired myocardial microcirculation, may help reduce cardiac mortality, including acute MI and sudden cardiac death, in hemodialysis patients. Consequently, oral nicorandil might improve survival rates in these patients by preventing cardiac death.^54^

Given the mixed results across the previously published literature for the effect of nicorandil on the outcomes apart from CIN in patients undergoing cardiac interventions, further research should be conducted to clarify the specific contexts in which nicorandil might be beneficial. This includes designing targeted clinical trials to assess its effects on major adverse events and other relevant outcomes. Updating clinical guidelines with the latest findings will help ensure that nicorandil is used effectively and appropriately.

Our study has several limitations. Firstly, the RCTs included open-label studies, which may increase the risk of bias. Secondly, the definition of CIN is based on serum creatinine changes, which can be influenced by several other factors. Thirdly, all the studies were conducted in Asian countries, except for one study in Egypt, which is a transcontinental country between Africa and Asia, making it uncertain whether the results are applicable elsewhere. Fourthly, the baseline renal functions varied among the participants in all the trials, which may explain the differential effect of nicorandil on CIN. Fifthly, the influences of cardiovascular comorbidities, medications, and contrast volume cannot be ignored. Sixthly, the patients included were only those with coronary heart disease undergoing PCI or CAG, so it is uncertain if the conclusions can be applied to other patients receiving contrast for different reasons. Seventhly, the trials used various dosages and forms of nicorandil, and there was insufficient data to conduct an analysis based on nicorandil dose along with dosage form. Long-term cardiovascular mortality also needs further exploration in future trials. Finally, we cannot rule out the potential impact of unpublished negative data on the real-world applicability of our findings.

## Conclusion

This meta-analysis concludes that nicorandil reduces the incidence of CIN after PCI or CAG without significantly increasing the risk of major adverse events including MI, cardiac death, heart failure, stroke, or dialysis. However, due to the small sample size and certain limitations, these findings should be confirmed through future well-designed, large-scale, double-blinded experimental studies before incorporating oral nicorandil into routine clinical care.

## Data Availability

All data produced in the present work are contained in the manuscript

## Statements and Declarations

### Financial support

No financial support was received for this study.

### Conflicts of interest

The authors report no relationships that could be construed as a conflict of interest.

## Acknowledgments

Not applicable.

## Availability of data

The data supporting this study’s findings are available from the corresponding author upon reasonable request.

---

Supplementary Figure 1: Risk of Bias in Included Studies

Supplementary Figure 2: Forest Plot of Major Adverse Events

Supplementary Figure 3: Forest Plot of Myocardial Infarction

Supplementary Figure 4: Forest Plot of Cardiac Death

Supplementary Figure 5: Forest Plot of Heart Failure

Supplementary Figure 6: Forest Plot of Stroke

Supplementary Figure 7: Forest Plot of Dialysis

## Notes

### Competing Interest Statement

The authors have declared no competing interest.

### Funding Statement

This study did not receive any funding

