## Supplementary Figure 1 for "Efficacy and Safety of Nicorandil for Prevention of Contrast-Induced Nephropathy in Patients Undergoing Coronary Procedures: A Systematic Review and Meta-Analysis"

| Study ID | D1 | D2 | D3 | D4 | D5 | Overall |
|---|---|---|---|---|---|---|
| Ali 2023 | + | + | + | + | + | + |
| Fan 2016 | + | + | + | + | ! | ! |
| Fan 2019 | ! | ! | + | + | + | ! |
| Iranirad 2017 | + | ! | + | + | ! | ! |
| Ko 2013 | + | ! | + | + | ! | ! |
| Moghaddam 2023 | + | + | + | - | - | - |
| Nawa 2015 | + | + | + | + | + | + |
| Otsuka 2021 | + | + | + | + | + | + |
| Yusuf 2024 | + | + | + | + | ! | ! |
| Zhang 2019 | + | + | + | + | ! | ! |
| Zhang 2020 | - | + | + | + | ! | - |
| Zhao 2017 | + | + | + | + | + | + |

+ Low risk

! Some concerns

- High risk

D1 Randomisation process

D2 Deviations from the intended interventions

D3 Missing outcome data

D4 Measurement of the outcome

D5 Selection of the reported result
