## Supplementary Figure 2 for "Efficacy and Safety of Nicorandil for Prevention of Contrast-Induced Nephropathy in Patients Undergoing Coronary Procedures: A Systematic Review and Meta-Analysis"

| | Nicorandil | | Control | | | Risk Ratio |
|---|---|---|---|---|---|---|
| Study or Subgroup | Events | Total | Events | Total | Weight | M-H, Random, 95% CI |
| Fan 2016 | 5 | 120 | 7 | 120 | 11.6% | 0.71 [0.23, 2.19] |
| Fan 2019 | 24 | 127 | 36 | 125 | 60.3% | 0.66 [0.42, 1.03] |
| Ko 2013 | 3 | 73 | 2 | 76 | 4.8% | 1.56 [0.27, 9.08] |
| Moghaddam 2023 | 6 | 172 | 0 | 172 | 1.8% | 13.00 [0.74, 228.99] |
| Zhang 2019 | 5 | 125 | 6 | 125 | 10.8% | 0.83 [0.26, 2.66] |
| Zhang 2020 | 5 | 150 | 6 | 150 | 10.7% | 0.83 [0.26, 2.67] |
| **Total (95% CI)** | | **767** | | **768** | **100.0%** | **0.77 [0.52, 1.13]** |
| Total events | 48 | | 57 | | | |

Heterogeneity: $Tau^2 = 0.01$; $Chi^2 = 5.18$, df = 5 (P = 0.39); $I^2 = 4\%$
Test for overall effect: Z = 1.34 (P = 0.18)

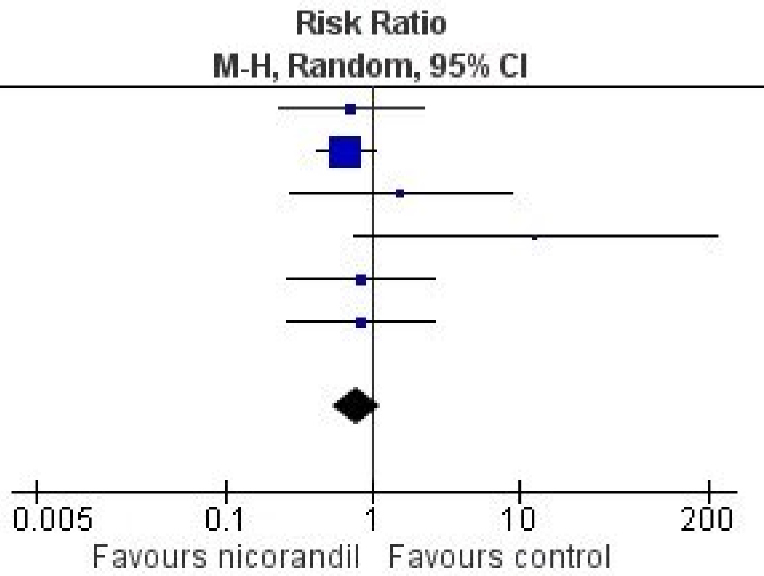
