## Supplementary Figure 3 for "Efficacy and Safety of Nicorandil for Prevention of Contrast-Induced Nephropathy in Patients Undergoing Coronary Procedures: A Systematic Review and Meta-Analysis"

| Study or Subgroup | Nicorandil Events | Nicorandil Total | Control Events | Control Total | Weight | Risk Ratio M-H, Random, 95% CI |
|---|---|---|---|---|---|---|
| Fan 2016 | 0 | 120 | 2 | 120 | 2.4% | 0.20 [0.01, 4.12] |
| Fan 2019 | 3 | 127 | 5 | 125 | 11.0% | 0.59 [0.14, 2.42] |
| Ko 2013 | 0 | 73 | 2 | 76 | 2.4% | 0.21 [0.01, 4.26] |
| Yusuf 2024 | 2 | 105 | 1 | 105 | 3.8% | 2.00 [0.18, 21.72] |
| Zhang 2019 | 23 | 125 | 23 | 125 | 80.3% | 1.00 [0.59, 1.69] |
| **Total (95% CI)** | | **550** | | **551** | **100.0%** | **0.90 [0.56, 1.43]** |
| Total events | 28 | | 33 | | | |

Heterogeneity: $Tau^2 = 0.00$; $Chi^2 = 2.83$, df = 4 (P = 0.59); $I^2 = 0\%$
Test for overall effect: Z = 0.45 (P = 0.65)

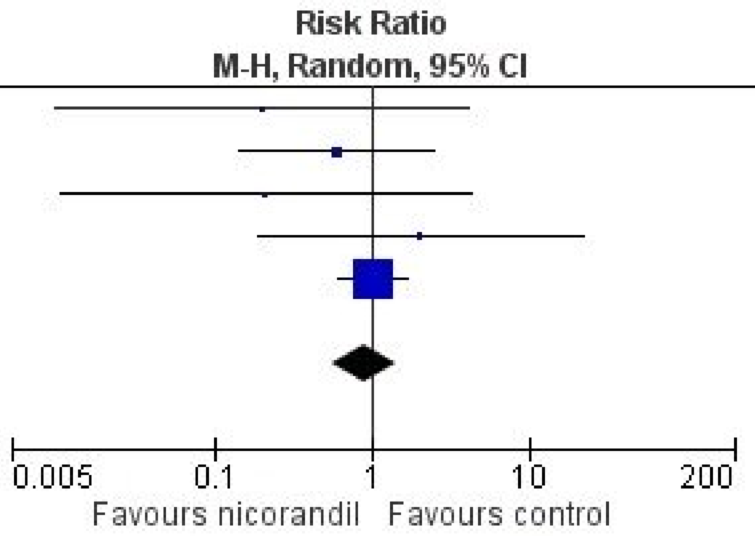
