## Supplementary Figure 4 for "Efficacy and Safety of Nicorandil for Prevention of Contrast-Induced Nephropathy in Patients Undergoing Coronary Procedures: A Systematic Review and Meta-Analysis"

| Study or Subgroup | Nicorandil Events | Nicorandil Total | Control Events | Control Total | Weight | Risk Ratio M-H, Random, 95% CI |
|---|---|---|---|---|---|---|
| Fan 2016 | 1 | 120 | 2 | 120 | 23.9% | 0.50 [0.05, 5.44] |
| Fan 2019 | 1 | 127 | 2 | 125 | 23.9% | 0.49 [0.05, 5.36] |
| Ko 2013 | 2 | 73 | 0 | 76 | 14.9% | 5.20 [0.25, 106.56] |
| Yusuf 2024 | 1 | 105 | 2 | 105 | 23.9% | 0.50 [0.05, 5.43] |
| Zhang 2019 | 1 | 125 | 0 | 125 | 13.4% | 3.00 [0.12, 72.94] |
| **Total (95% CI)** | | **550** | | **551** | **100.0%** | **0.90 [0.28, 2.88]** |
| Total events | 6 | | 6 | | | |

Heterogeneity: $Tau^2 = 0.00$; $Chi^2 = 2.59$, df = 4 (P = 0.63); $I^2 = 0\%$

Test for overall effect: Z = 0.18 (P = 0.86)

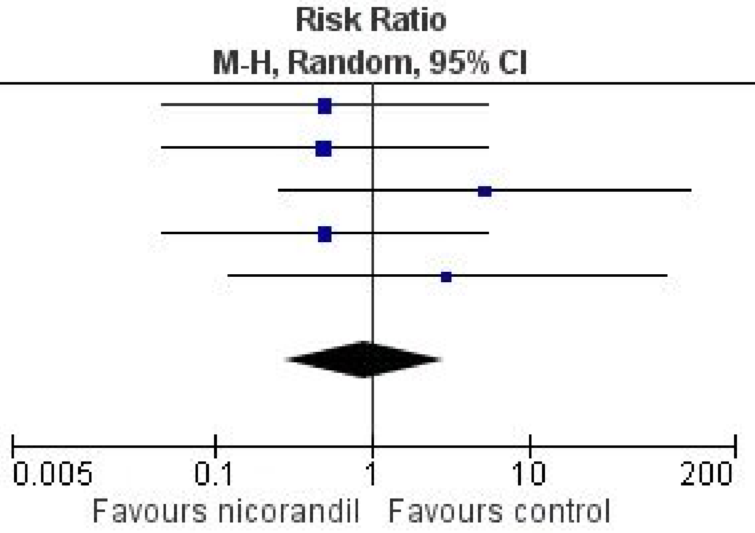
