## Supplementary Figure 5 for "Efficacy and Safety of Nicorandil for Prevention of Contrast-Induced Nephropathy in Patients Undergoing Coronary Procedures: A Systematic Review and Meta-Analysis"

| Study or Subgroup | Nicorandil Events | Nicorandil Total | Control Events | Control Total | Weight | Risk Ratio M-H, Random, 95% CI |
|---|---|---|---|---|---|---|
| Fan 2016 | 2 | 120 | 3 | 120 | 16.9% | 0.67 [0.11, 3.92] |
| Fan 2019 | 5 | 127 | 5 | 125 | 35.8% | 0.98 [0.29, 3.32] |
| Yusuf 2024 | 2 | 105 | 3 | 105 | 16.9% | 0.67 [0.11, 3.91] |
| Zhang 2019 | 3 | 125 | 3 | 125 | 21.2% | 1.00 [0.21, 4.86] |
| Zhang 2020 | 1 | 150 | 2 | 150 | 9.3% | 0.50 [0.05, 5.46] |
| **Total (95% CI)** | | **627** | | **625** | **100.0%** | **0.81 [0.39, 1.68]** |
| Total events | 13 | | 16 | | | |

Heterogeneity: $Tau^2 = 0.00$; $Chi^2 = 0.42$, df = 4 (P = 0.98); $I^2 = 0\%$

Test for overall effect: Z = 0.56 (P = 0.58)

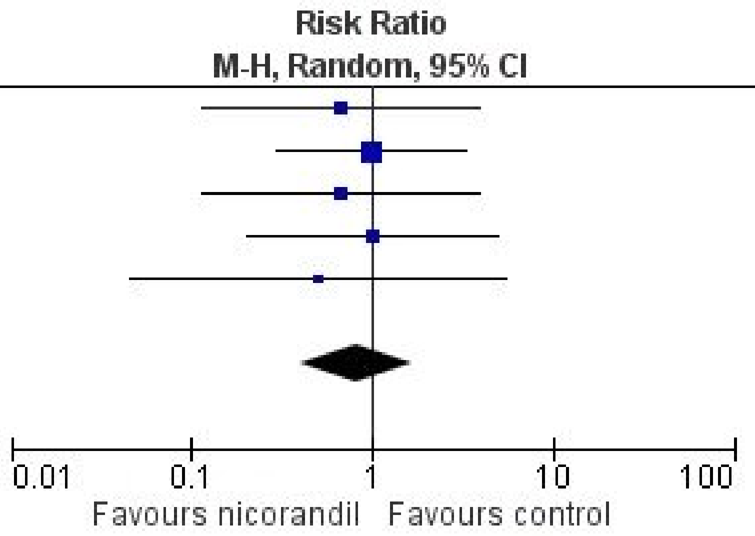
