## Supplementary Figure 6 for "Efficacy and Safety of Nicorandil for Prevention of Contrast-Induced Nephropathy in Patients Undergoing Coronary Procedures: A Systematic Review and Meta-Analysis"

| | Nicorandil | | Control | | | Risk Ratio |
|---|---|---|---|---|---|---|
| Study or Subgroup | Events | Total | Events | Total | Weight | M-H, Random, 95% CI |
| Fan 2016 | 1 | 120 | 0 | 120 | 10.4% | 3.00 [0.12, 72.91] |
| Fan 2019 | 4 | 127 | 6 | 125 | 68.8% | 0.66 [0.19, 2.27] |
| Zhang 2019 | 1 | 125 | 0 | 125 | 10.4% | 3.00 [0.12, 72.94] |
| Zhang 2020 | 1 | 150 | 0 | 150 | 10.4% | 3.00 [0.12, 73.06] |
| **Total (95% CI)** | | **522** | | **520** | **100.0%** | **1.05 [0.38, 2.95]** |
| Total events | 7 | | 6 | | | |

Heterogeneity: Tau² = 0.00; Chi² = 1.81, df = 3 (P = 0.61); I² = 0%

Test for overall effect: Z = 0.10 (P = 0.92)

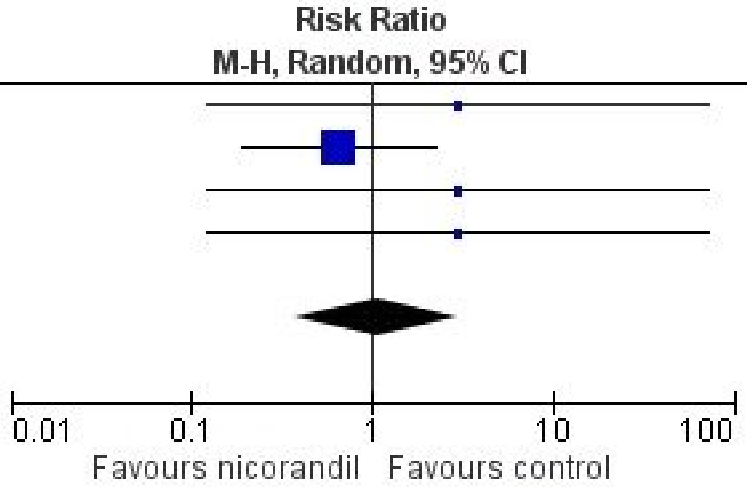
